# Systematic Review of the Diagnostic Imaging Evaluation of Pulsatile Tinnitus

**DOI:** 10.1101/2025.02.25.25322858

**Authors:** Meghan P. Jairam, Simon Kidanemariam, Aleena Malik, C. Eduardo Corrales, Chong Hyun Suh, Jeffrey P Guenette

## Abstract

**Objective:** Aggregate published data on the imaging of pulsatile tinnitus as a step toward building a framework for an evidence-based approach to diagnostic imaging for this symptom.

**Materials & Methods:** A systematic review was performed. PUBMED and EMBASE were searched on December 1, 2021 for English-language articles on diagnostic imaging of pulsatile tinnitus. Articles that involved non-standard imaging techniques and those that focused on management of pulsatile tinnitus were excluded. Extracted data included: number of males and females; signs, symptoms, and physical examination findings with associated patient counts; imaging findings; count of patients with imaging-identified cause of pulsatile tinnitus; reported associated interventions and outcomes.

**Results:** 41 articles were included with a total of 2,633 reported patients. 10 studies were prospective. MRA appears to be capable of identifying many of the same pathologies traditionally diagnosed with DSA. Few head-to-head comparisons were performed. In head-to-head comparisons of MRI and MRA, MRA was often able to identify more pathology. There was no clear relationship identified between specific symptoms and the imaging modality chosen, indicating that the imaging evaluation of pulsatile tinnitus is likely sensitive to the preferences of the evaluating provider.

**Conclusion:** There is limited evidence to inform best practices for the initial imaging evaluation of pulsatile tinnitus and preference-sensitive provider decisions will continue to guide the pulsatile tinnitus workup. We encourage prospective studies with multimodality imaging comparisons to build evidence that would support the development of more effective, efficient, and equitable protocols and pathways for the imaging evaluation of pulsatile tinnitus.

**Clinical Relevance Statement:** Evidence is not available in support of any single optimal imaging evaluation of pulsatile tinnitus, but prudent imaging will include evaluation for life-threatening causes such as dural arteriovenous fistula and arteriovenous malformation.

**Key Points:**

- Many imaging findings associated with pulsatile tinnitus may not be causal.
- Current evidence is insufficient to indicate the optimal imaging evaluation.
- Local preference-based imaging algorithms are reasonable in the absence of new high-level evidence.

## Introduction

Pulsatile tinnitus is a rhythmic sound, often described by patients as whooshing or thumping, that is heard in either one or both ears.[1] Pulsatile tinnitus can be categorized as objective when the examining physician can hear the rhythmic sound or subjective when the sound is only discernable to the patient.[2] To some patients, the constant rhythmic sound can be extremely debilitating and can lead to anxiety, depression, and a lower quality of life.[3] Furthermore, pulsatile tinnitus can be the sign of a serious underlying condition, such as arteriovenous fistula,[4] which can have a mortality rate as high as 20% when left untreated.[5] Almost half of pulsatile tinnitus cases ultimately have a treatable cause.[6, 7]

Patients presenting with pulsatile tinnitus-like symptoms typically undergo a physical examination and diagnostic imaging workup. In approximately 70% of cases, imaging alone can help identify underlying etiologies.[8] However, arterial, venous, neoplastic, middle and inner ear, and neurological causes of pulsatile tinnitus are best identified on specific types of imaging examinations (e.g. temporal bone CT versus MRA) and may not be identifiable on others. With multiple potential imaging examinations to choose from and multiple published diagnostic algorithms, selecting the proper imaging examination can be challenging.[8–11] There is not currently aggregated data on the relative utility of the various possible imaging examinations, allowing for substantial preference-sensitive variation in examination ordering patterns based on individual provider’s familiarity and anecdotal experience with pulsatile tinnitus.

The goal of this systematic review is to aggregate published data on the imaging of pulsatile tinnitus as a step toward building a comprehensive framework for an evidence-based approach to diagnostic imaging for this symptom. We include presenting symptoms, physical examination findings, imaging modality chosen based on symptoms, and the full range of associated diagnostic findings for each published study of each imaging examination type.

## Methods

This systematic review of published literature did not require IRB approval and was reported in accordance with Preferred Reporting Items for Systematic reviews and Meta-Analyses (PRISMA) guidelines.[12, 13] Given the expectation of minimal evidence for analysis, a review protocol was not prepared or registered. PUBMED and EMBASE were searched from database inception to December 1, 2021 inclusively for English-language articles.

The search term used was ("pulsatile tinnitus") AND ((computed tomography) OR (CT) OR (CT angiography) OR (CTA) OR (magnetic resonance) OR (MRI) OR (MR angiography) OR (MRA) OR (ultrasound) OR (sonograph*) OR (angiography)). For Level 1 phase of screening, 2 reviewers (MPJ, RC) independently screened all articles by their title and abstract and excluded articles that did not report on signs/symptoms suspicious of pulsatile tinnitus prior to imaging work-up. Articles detailing the evaluation or imaging of non-pulsatile tinnitus were excluded.

Additionally, case reports, case series with fewer than three patients, review articles, letters, comments, notes, and editorials were excluded. In the event of conflict, discussion between the reviewers was led to consensus or, if needed, a third reviewer (JPG) was involved to resolve the conflict.

The remaining studies underwent Level 2 screening where full texts were assessed to determine eligibility by two independent reviewers (MPJ, SK). In the event of conflict, discussion between the reviewers was led to consensus or, if needed, a third reviewer (JPG) was involved to resolve the conflict. Specifically, those articles with the following imaging modalities were included: Head CT, Temporal Bone CT, Head/Neck CTA, Brain MRI, Temporal Bone MRI (also called Internal Auditory Canal MRI), Head/Neck MRA/MRV, Ultrasound (US), digital subtraction angiography (DSA), all with standard techniques. At this stage, articles that involved non-standard imaging techniques and those that focused on management of pulsatile tinnitus, as opposed to the initial work-up, were excluded.

The remaining eligible studies underwent a final round of data extraction and synthesis for inclusion in this systematic review. Extracted data included: number of males and females; signs, symptoms, and physical examination findings with associated patient counts; imaging findings; count of patients with imaging-identified cause of pulsatile tinnitus; reported associated interventions and outcomes. Bias was assessed by one author (JPG) using the Revised Risk of Bias Assessment Tool for Nonrandomized Studies of Interventions (RoBANS 2).[14]

## Results

A total of 670 articles were identified from database search. Level 1 screening was performed on 605 articles after duplicates were removed, resulting in 77 articles for Level 2 full-text review. The Level 2 screening yielded 41 articles for inclusion.[6, 8, 15–53] A PRISMA flow diagram is provided as Figure 1. Supplementary Tables 1 through 7 list the extracted details from the included articles.

**Figure 1:**
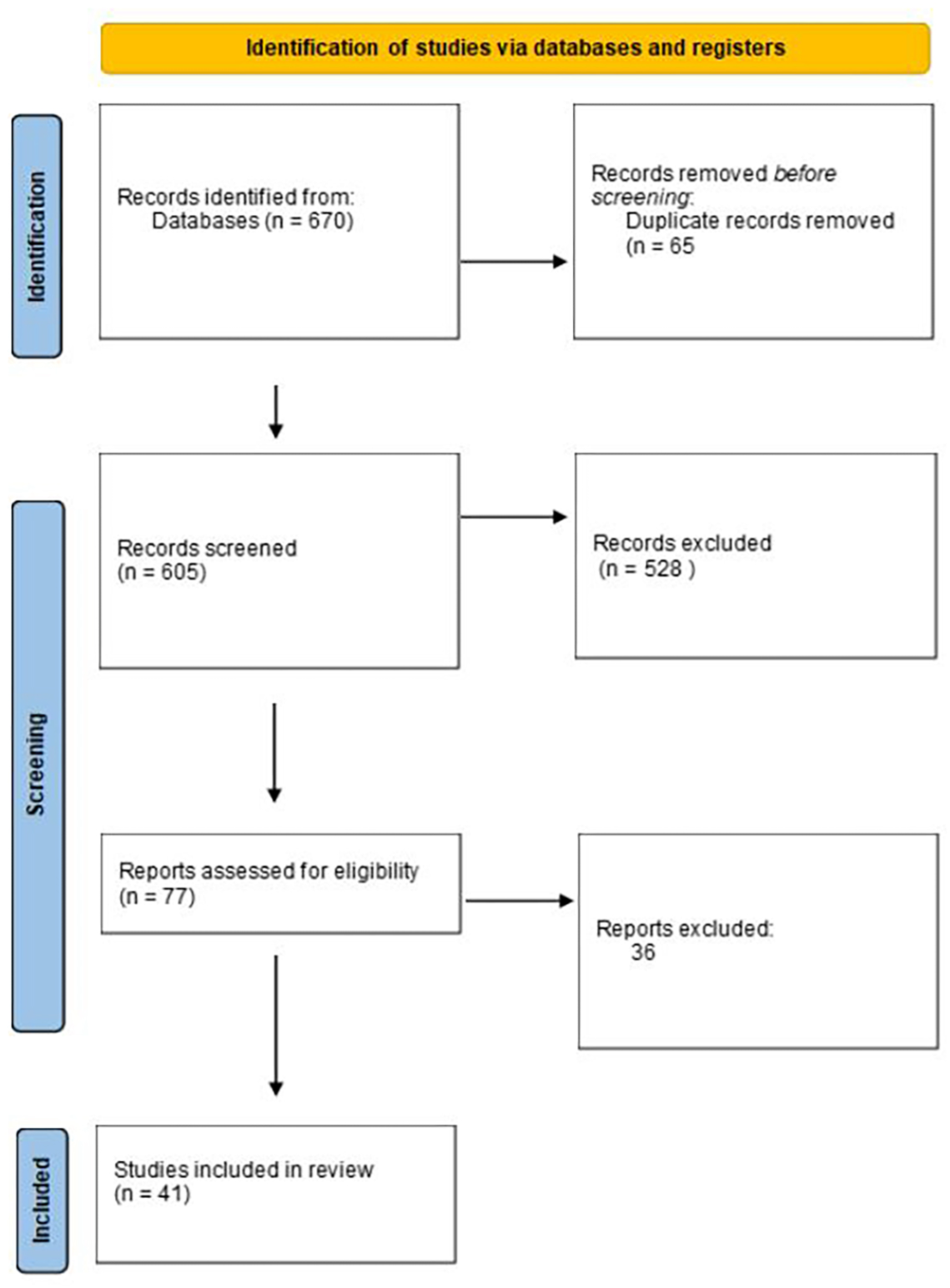
PRISMA Flow Diagram

The 44 studies included a total of 2,633 patients. In many studies, the number of patients who had imaging with a specific imaging modality was not specified. Of the studies in which numbers were specified, 401 underwent Head CT, an unspecified total number underwent Temporal Bone CT and MRA, 143 Head/Neck CTA/CTV, 346 Brain MRI, zero Temporal Bone MRI, 120 Head/Neck MRA, 369 Ultrasound (US), 290 digital subtraction angiography (DSA), and 149 catheter angiography not specified as DSA. High risk of bias was present in most studies, particularly with regard to comparability of the target group (no comparison group in most studies), blinding of assessors, and selective outcome report (Supplemental Table 8).

Ten studies were prospective studies.[20, 27, 30, 38, 43, 44, 46, 47, 50, 51] Varied imaging modalities were used to evaluate pulsatile tinnitus in these prospective studies, including DSA, CTA/CTV, CT, MRA, MRI, and carotid doppler ultrasound.

In addition to pulsatile tinnitus, 83 patients also presented with dizziness, 255 patients with headache, and 41 patients with hearing loss, however there is likely bias based on the assessing provider speciality. Rarely, studied patients presented with hypertension, head trauma, dysphasia, aphasia, papilledema, amaurosis fugax, hemiparesis, or Horner’s syndrome in addition to pulsatile tinnitus.

### Physical examination findings and type of imaging

A variety of imaging modalities were used to workup the disappearance of pulsatile tinnitus upon compression of the ipsilateral jugular vein.

For example, in those patients with the disappearance of pulsatile tinnitus upon compression of the ipsilateral vein, temporal bone CT [18, 19] was used to diagnose high jugular bulb. CT was used to diagnose sigmoid sinus diverticulum, jugular bulb diverticulum, dehiscent jugular bulb, large emissary vein, sinus thrombosis, and petrosquamosal sinus.[25] CTA was used to diagnose dominant venous systems, transverse sinus diverticulum/stenosis, high riding jugular bulb without bone dehiscence, IJV stenosis, intra mastoid venous channel, external carotid artery stenosis and other arterial and venous causes, in addition to focal defects of the mastoid shelf. [8, 34, 53, 54]

In other patients with disappearance of pulsatile tinnitus upon ipsilateral compression of the vein, MRA/V was used to diagnose transverse sigmoid sinus stenosis[55] as well as jugular bulb diverticulum, large condylar vein, prominent emissary vein, venous sinus aneurysm, venous sinus stenosis, dural arteriovenous fistula, and arteriovenous malformation.[49] Digital subtraction was used to diagnose dural arteriovenous fistula, fibromuscular dysplasia, sigmoid sinus diverticulum, venous outflow obstruction, and mastoid emissary vein.[31, 33]

In patients with an ipsilateral carotid bruit and no otoscopy findings, carotid ultrasonography was ordered.[22] In this study, among patients who underwent carotid ultrasonography, only 4/34 patients obtained an imaging diagnosis and were found to have atherosclerosis.

In those patients with conductive and sensorineural hearing loss, some providers obtained a temporal bone CT, and those patients were found to have a jugular bulb diverticulum and vestibular aqueduct dehiscence.[56] Other providers obtained a CTA/V and those patients were found to have extracranial tortuosities of the ICA.[5]

Some patients were found to have a bruit and/or observed tumor with an occasional red hemotympanum on physical examination. In these patients, catheter angiography was used to identify jugular glomus, AV malformation, external carotid artery stenosis, vertebrojugular fistula, glomus tympanicum, acoustic neuroma.[28, 57] In other patients (some of which had a red mass on physical examination), digital subtraction angiography was used to identify a vascular mass, frontal lobe arteriovenous malformation, jugular vein thrombosis, or large jugular bulb.[58] In Sila,[42] several patients had audible bruits and digital subtraction angiography was used to identify arteriovenous malformations, ICA occlusion/stenosis, carotid dissections, and arterial ectasia.

### Sources of pulsatile tinnitus and type of imaging

All 7 studies using ultrasound evaluated for and showed atherosclerosis but did not show any other causes of pulsatile tinnitus.

Although catheter angiography is considered one of the gold standards, MRA/MRV was used to identify the same pathologies.

### Direct comparison of types of imaging

When comparing imaging modalities to one another for the identification of specific causes of pulsatile tinnitus, carotid, doppler and cranial sonography appear adequate at evaluating specifically for atherosclerosis as a cause of pulsatile tinnitus.

In head-to-head comparisons of MRI and MRA, MRA was often able to identify more pathology. For example, in Dietz et al. 1994, when compared with MRI, MRA better identified dural arteriorvenous fistula, high jugular bulb, and jugular bulb diverticulum.[24]

In Tsai et al. 2016, out of 28 patients with suspicion for dural arteriovenous fistula based on initial imaging with carotid duplex sonography, 25 underwent additional imaging with either MRI/MRA or CT/CTA, suggesting that if the suspicion for a dural arteriovenous fistula is high enough, an MRI/MRA or CT/CTA should be obtained.[59] Specifically, the MRI/MRA features of dural arteriovenous fistula included cerebral sinus opacification, enlargement or clustering of arterial external carotid artery branches, and abnormal vasculature around the brain surface. CT/CTA revealed vessels close to or inside skull bones. The authors report that if all the patients in this study (n = 155) initially underwent MRI and MRA, the total cost would have been $154,070. Because the patients were initially screened with carotid duplex sonography, the costs were significantly lower.

## Discussion

In this systematic review, we aggregated all published research data on the diagnostic imaging of pulsatile tinnitus. We outlined all reported presenting symptoms, physical examination findings, imaging modality chosen based on symptoms, and the full range of associated diagnostic findings for each published study of each imaging examination type. There was no clear relationship identified between specific symptoms and the imaging modality chosen, indicating that the imaging evaluation of pulsatile tinnitus is largely sensitive to the preferences of the evaluating provider.

The most common finding on physical examination was the disappearance of pulsatile tinnitus upon compression of the ipsilateral jugular vein. For this physical examination finding, a variety of imaging modalities were used in diagnosis. To evaluate specifically for atherosclerosis as a cause of pulsatile tinnitus, carotid, doppler and cranial sonography were determined to be equally useful. In the published English-language literature reviewed, very few head-to-head comparisons were performed, except for head-to-head comparisons of MRI and MRA, in which MRA was often able to identify more pathology.[24] MRA appears to be capable of identifying many of the same pathologies traditionally diagnosed with DSA.

There are several limitations of this study. First, the literature included in this review consisted mostly of retrospective studies with high risk of bias. Some potential causes of pulsatile tinnitus, such as superior semicircular canal dehiscence, are known but there were no diagnostic imaging studies on the topic that were identified in our screening process.[60, 61] Second, there were very few prospective and head-to-head comparison studies. There was no suitable quantitative data to perform a meta-analysis.

In conclusion, there is limited evidence to inform best practices for the initial imaging evaluation of pulsatile tinnitus and preference-sensitive provider decisions will continue to guide the pulsatile tinnitus workup. We encourage prospective studies with multimodality imaging comparisons to build evidence that would support the development of more effective, efficient, and equitable protocols and pathways for the imaging evaluation of pulsatile tinnitus.

## Data Availability

All data produced in the present work are contained in the manuscript

## Abbreviations

CT: X-Ray Computed Tomography
CTA: X-Ray Computed Tomography Angiography
MRI: Magnetic Resonance Imaging
MRA: Magnetic Resonance Angiography
MRV: Magnetic Resonance Venography
US: Ultrasound
DSA: Digital Subtraction Angiography

---

**Supplemental Table 1.** Carotid, doppler, and cranial ultrasonography.

| <u>Study</u> | <u>Gender: F, M</u> | <u>Signs, symptoms, physical exam</u> | <u>Doppler ultrasonography findings</u> | <u>Proportion of patients with imaging-identified pulsatile tinnitus</u> | <u>Interventions/Outcomes</u> |
| --- | --- | --- | --- | --- | --- |
| Bae et al. 2015 | 47, 10 | -Suspected arterial origin of PT | Atherosclerosis | 57/57 (total patients, unspecified number obtained each modality of imaging) |  |
| Daneshi et al. 2004<sup>22</sup> | 20, 14 | - 10 with objective PT<br>- 24 with subjective PT<br>- 3 with bilateral PT<br>All 34 with ipsilateral carotid bruit and normal otoscopy | Atherosclerosis | 4/34 | N/A |
| Waldvogel et al. 1998<sup>52</sup> | Unspecified | - 35 with objective PT<br>- 28 with subjective PT<br>- 1 with facial spasm | -11/11 with increased flow velocity in external carotid and occipital arteries<br>- 7/12 with atherosclerotic stenosis<br>-5/12 with unspecified ultrasound findings. | 11/16 (16 underwent cranial ultrasound)<br>12/40 (40 with vascular pathology underwent carotid ultrasound) | N/A |
| Tsai et al. 2016<sup>59</sup> | Unspecified | -155 with PT | -25/28: suspicion for DAVF, underwent further imaging with MR/MRA or CT/CTA<br>-3/28 carotid stenosis, no further imaging | 28/155 | N/A |
| Terzi et al. 2015<sup>62</sup> | 22, 12 | -17 with right-sided PT<br>-11 with left-sided PT<br>-6 with bilateral PT | Not specified | Not specified | N/A |
| Sismanis, Smoker 1994<sup>46</sup> | Unspecified. Out of the 15 with DA, 11 women 4 men | Used in patients suspected with ACAD | Moderate to severe CA stenosis | 15/100 | N/a |
| Sismanis et al. 1994<sup>47</sup> | 8, 4 | -5 left PT<br>-1 bilateral<br>-6 right PT | Mild to severe atherosclerosis in the ICA and CCA | 12/12 | N/A |
| Sonmez et al. 2007<sup>48</sup> | 30, 44 | -40 right PT<br>-28 left PT<br>12 bilateral PT | Atherosclerotic changes were found in different levels and degrees of the extracranial carotid arteries in 16 patients by Doppler studies. | 50/74, 12 with doppler ultrasonography | N/A |

**Supplemental Table 2.** Temporal bone CT and CT.

| <u>Study</u> | <u>Gender: F, M</u> | <u>Signs, symptoms, physical exam</u> | <u>Temporal Bone CT or CT findings</u> | <u>Proportion of patients with imaging-identified pulsatile tinnitus</u> | <u>Interventions/Outcomes</u> |
| --- | --- | --- | --- | --- | --- |
| Bae et al. 2015<sup>16</sup> | 47, 10 | -Tympanic mass medial to eardrum | Jugular bulb anomaly<br>Glomus tumor | 57/57 |  |
| Berguer et al. 2015<sup>18</sup> | 7,0 | -7 unilateral<br>-Pt disappears upon compression of the ipsilateral jugular vein | High jugular bulb | 7/7 | -5 patients had ligation performed above the level of the facial vein, with complete resolution of symptoms.<br>-The other 2 patients had ligation below the facial vein and experienced a decrease of pulsatile tinnitus. |
| Buckwalter et al. 1983<sup>19</sup> | 3,0 | -3 unilateral<br>-Disappearance of pt symptoms with ipsilateral jugular compression<br>-In 2/3 patients, symptoms worsened with exercise | High jugular bulb | 3/3 | -2/3 opted for IJV ligation, resulting in immediate resolution of symptoms post-op |
| Dong et al. 2015<sup>25</sup> | 215,27 | -83 pts had dizziness,<br>-26 had a headache<br>-Hearing loss in 34 patients<br>-Pt symptoms resolution after compression of ipsilateral IJV | -Dehiscent sigmoid plate<br>-lateral sinus stenosis<br>- high jugular bulb<br>- sigmoid sinus diverticulum<br>- jugular bulb diverticulum<br>- dehiscent jugular bulb<br>- large emissary vein<br>- sinus thrombosis<br>- petrosquamosal sinus | 228/242 had at least one anatomic abnormality on the symptomatic side of CT | N/A |
| Friedmann et al. 2010<sup>56f</sup> | 1,2 | -1 conductive hearing loss<br>-1 sensorineural hearing loss<br>-2 had bilateral pt and 1 had unilateral | -2/3 jugular bulb diverticulum<br>-1/3 jugular bulb<br>- 3/3 with vestibular aqueduct dehiscence on CT | 3/3 | N/A |
| Kline et al. 2020<sup>29</sup> | 21,1 | -MRI and MRV were performed initially to confirm diagnosis of IIH | -Sigmoid sinus wall abnormalities | 7/10 pts with PT had imaging findings | 2/14 subjects experienced temporary relief in symptoms after LP |
| Lynch et al. 2021<sup>6</sup> | 186, 65 | Unspecified | -Venopathy<br>-Middle/inner ear pathology<br>-Neoplasm | 32/53 | Lynch et al. 2021<sup>6</sup> |
| Sonmez et al. 2007<sup>48</sup> | 30,44 | -30 patients were hypertensive | -21 high JV bulb<br>- 16 atherosclerosis<br>-3 dehiscent jugular bulb | 50/74 had tinnitus causing lesions on HRCT | N/A |
|  |  |  | -1 jugular diverticulum<br>-2 Dural AVF<br>- 1 aberrant ICA<br>-2 glomus tympanicum<br>- 3 aneurysm of ICA<br>-1 patient with intracranial tumor |  |  |
| Sismanis et al. 1990<sup>63</sup> | 30 total, unspecified | Presented with tinnitus, suspicion for benign intracranial hypertension. | All findings made on both CT and MRI. 12 patients with empty sella, 4 patients with small ventricles, and Chiari Type 1 malformation in one patient. | 17/30 | Final diagnosis made based on opening LP pressures |
| Sismanis et al. 1998<sup>43</sup> | 145 total, unspecified | Retrotympanic mass | Glomus tympanicum<br>Aberrant carotid artery<br>Jugular bulb abnormality | Unspecified out of a total of 145 patients |  |

**Supplemental Table 3.** Catheter angiography.

| <u>Study</u> | <u>Gender: F, M</u> | <u>Signs, symptoms, physical exam</u> | <u>Angiography Findings</u> | <u>Proportion of patients with imaging-identified pulsatile tinnitus</u> | <u>Interventions/Outcomes</u> |
| --- | --- | --- | --- | --- | --- |
| Buckwalter et al. 1983<sup>19</sup> | 3,0 | -3 unilateral<br>-Disappearance of pt symptoms with ipsilateral jugular compression<br>-In 2/3 patients, symptoms worsened with exercise | Enlarged jugular bulb | 3/3 | -2/3 opted for IJV ligation, resulting in immediate resolution of symptoms post-op |
| Harris et al. 1979<sup>57</sup> | N/A | -Audible murmur in 9/15<br>-Observed tumor on otoscopy in 3 patients | -3 Jugular Glomus<br>- 8 AV Malformation<br>-1 External Carotid Artery Stenosis | 12/15 | -Patients with jugular glomus tumor and AV malformation underwent gelatin sponge embolization<br>-Varying amounts of symptom reduction and symptom elimination were observed |
| Holgate et al. 1977<sup>28</sup> | 4,4 | -Bruit auscultated<br>- red hemotympanum<br>- hearing loss<br>- contralateral carotid occlusion<br>-carotid body tumor | -2 AVM<br>-1 Vertebro-jugular Fistula<br>-1 Glomus Tympanicum<br>-1 Glomus Jugulare Tumor<br>-1 Acoustic Neuroma<br>-1 Primitive Hypoglossal Artery<br>-1 Chiari malformation | 8/8 | -Out of 6 who underwent surgery, 4 had resolution, 1 had reduction, and 1 developed continuous PT |
| Pelkonen et al. 2004<sup>36</sup> | 9,7 | -6/16 had dysphasia, aphasia, amaurosis fugax, and hemiparesis<br>-4/16 Horner's syndrome<br>-2/16 vertigo and dysgeusia<br>-13/16 headache/neckache | -11 Arterial dissection involved unilateral ICA<br>-2 bilateral ICA dissection<br>-2 unilateral vertebral artery dissection<br>-1 patient with both bilateral ICA and bilateral VA dissection. | 16/16 | -14/16 patients received anticoagulant therapy<br>-1 patient had ligation of the ICA to treat pseudoaneurysm<br>- In majority of the patients, PT symptoms subsided |
| Russell et al. 1995<sup>37</sup> | 4, 0 | -4/4 audible bruit | -Focal narrowing of transverse dural sinus | 4/4 |  |

**Supplemental Table 4.** CTA/V.

| <u>Study</u> | <u>Gender F, M</u> | <u>Signs, symptoms, physical exam</u> | <u>CTA/V findings</u> | <u>Proportion of patients with imaging-identified pulsatile tinnitus</u> | <u>Interventions/Outcomes</u> |
| --- | --- | --- | --- | --- | --- |
| Bae et al. 2015<sup>16</sup> | 47, 10 | -Suspected arterial origin of PT | Dural AV fistula<br>Intracranial aneurysm | 57/57 (total patients, unspecified number obtained each modality of imaging) |  |
| Krishnan et al. 2006<sup>54</sup> | 9, 7 | -Decrease in PT symptoms upon compression of ipsilateral neck | -6 dominant venous systems<br>-1 transverse sinus diverticulum<br>-1 transverse sinus stenosis<br>-1 high riding jugular bulb without bone dehiscence<br>-1 external carotid artery stenosis | 7/16 | -One patient had surgery for right transverse sinus diverticulum followed by resolution of symptoms |
| Lynch et al. 2021<sup>6</sup> | 186, 65 | Unspecified | -Arteriopathy | 21/30 |  |
| Mattox et al., 2008<sup>8</sup> | 38,16 | -Compression of ipsilateral jugular vein or carotid artery (only for younger patients) | - 14 were considered arterial<br>-23 venous<br>- 15 were indeterminate in origin | 39/54 | N/A |
| Mundada et al. 2015<sup>34</sup> | 20,12 | 14/32 experienced a decrease in their symptoms after compression of the ipsilateral jugular vein | -12 dehiscent sigmoid plate (DSP)<br>-2 DSP with sigmoid sinus diverticulum<br>- 13 IJV stenosis<br>-3 high jugular bulb<br>-3 Dehiscent jugular bulb<br>- 2 arge posterior condylar vein<br>-1 Large posterior emissary vein from the transverse sinus<br>-1 Intra mastoid venous channel -1 Transverse sinus stenosis -1Sigmoid sinus stenosis<br>-1 Jugular bulb stenosis | 31/32 | N/A |
| Otto et al. 2007<sup>64</sup> | 3,2 | 2/5 had audible bruit on the symptomatic side<br>3/5 had a reduction in pt after compression of ipsilateral IJV<br>2/5 sensorineural hearing loss | -sigmoid sinus diverticulum | 5/5 | -3/5 underwent surgery for transmastoid reconstruction for SSD and all had resolution of symptoms |
| Sismanis et al. 2008<sup>45</sup> | 3,0 | Audiogram revealed bilateral mild high frequency sensorineural hearing loss in 1 patient | -extracranial tortuosities of ICA | 3/3 | N/A |
| Tsai et al. 2016<sup>59</sup> | Unspecified | -155 with PT | -25/28: suspicion for DAVF, underwent further imaging with MR/MRA or CT/CTA<br>-3/28 carotid stenosis | 28/155 | N/A |
| Xue et al., 2012<sup>53</sup> | N/A | 2 out of the 3 patients eliminated bruit by compressing ipsilateral upper neck | -focal defects of the mastoid bone shelf | 3/3 | Transmastoid reconstruction of the mastoid bone shell overlying the transverse-sigmoid sinus with resolution of symptoms |

**Supplemental Table 5.** MRA/V.

| <u>Study</u> | <u>Gender: F, M</u> | <u>Signs, symptoms, physical exam</u> | <u>MRA/ V Findings</u> | <u>Proportion of patients with imaging-identified pulsatile tinnitus</u> | <u>Interventions/Outcomes</u> |
| --- | --- | --- | --- | --- | --- |
| Bae et al. 2015<sup>16</sup> | 47, 10 | -Suspected arterial origin of PT | Dural AV fistula<br>Intracranial aneurysm | 57/57 (total patients, unspecified number obtained each modality of imaging) |  |
| Baomin 2014<sup>55</sup> | 44,2 | -44 unilateral (34 right-sided, 12 left-sided)<br>-2 bilateral<br>Pulsatile tinnitus symptoms reduced with head rotation or ipsilateral neck compression | -20/46 with transverse sigmoid sinus stenosis. The others were identified by DSA. | 20/46 | -All patients failed an unspecified medical therapy.<br>-All patients underwent angioplasty and stenting and had complete resolution with no recurrent symptoms after 36 moth follow-up period |
| Tao 2021<sup>49</sup> | 139, 48 | -20 bilateral PT<br>-41 left PT<br>-60 right PT<br>-22 migraine<br>-9 head trauma history<br>-63 headaches<br>-36 visual symptoms | Venous sinus stenosis, post-stenotic venous aneurysm/ diverticulum, jugular bulb diverticulum, DAVF, carotid cavernous fistula | 105 | N/A |
| Amans 2018<sup>65</sup> | 25, unspecified gender | PT suspected to be of venous etiology (improved with neck compression) | Sigmoid Sinus Diverticulum | 5 |  |
| Lynch et al. 2021<sup>6</sup> | 186, 65 | Unspecified | -Arteriopathy<br>-Venopathy<br>-Neoplasm | 30/65 |  |
| Lyu 2018<sup>32</sup> | 35, 14 | -39 right ear<br>-10 left ear | MRA was used as key imaging in diagnosis of ICA stenosis, DAVF | 49 | N/A |
| Waldvogel 1998<sup>52</sup> | 58, 26 | n/a | Value of MRA cannot be evaluated because most patients were seen before using this imaging method | 57, only 5 by MRA | N/A |
| Dietz 1994<sup>24</sup> | 31, 17 | -21 right PT<br>-4 bilateral PT<br>-18 left PT | DAVF, paraganglioma, extracranial AVF, high jugular bulb, jugular bulb diverticulum, stenosis of transverse sinus (only identified on MRA), aberrant ICA, pial AVM (only identified on MRA), stenosis of ICA (only identified on MRA), carotid dissection (only identified on MRA), tortuous ICA (only identified on MRA), maxillary AVM (only identified on MRA) | 28 | N/A |
| Sanchez 1998<sup>66</sup> | 15, 1 | -6 right PT<br>-5 left PT<br>-4 bilateral PT<br>-2 head PT | -4 hypoplasia of artery<br>-5 basilar artery<br>-2 intracranial aneurysms<br>-1 intracranial hypertension | 13 | N/A |
| Shin 2000<sup>39</sup> | 32, 22<br>Second review:<br>20, 13 | Second review:<br>-right PT in 11<br>-bilateral in 5<br>-left in 17 | high riding jugular bulb (9),<br>dominant transverse sinus (5),<br>attenuated transverse sinus (3),<br>fibromuscular dysplasia (1), carotid<br>dissection healing (1). Serous otitis<br>media (4), TS DAVD (3).<br>Catheter angiography was used as<br>the gold standard in this review. | 3 by MRA, then 33 more in<br>second review | N/A |
| Shweel 2013<sup>41</sup> | 11, 16 | -right 13<br>-left 18<br>-4 bilateral | DAVF (4), high jugular bulb (2),<br>aneurysm of ICA (1), aberrant ICA<br>(1), vertebral artery hypoplasia (2),<br>glomus tumor (1) | 11 | N/A |
| Tsai 2016<sup>59</sup> | n/a | N/a | DAVF, high jugular bulb, tortuous<br>ICA or VA, DAVF, MCA stenosis,<br>subclavian artery stenosis | 77 | N/A |
| Sismanis et al.1994<sup>46</sup> | n/a | MRA was used after PT was confirmed<br>by CA and only on selected patients | Tortuous basilar artery<br>AVM | 12/100 MRA was used.<br>Confirmed AVM in 3. | N/A |

**Supplemental Table 6.** MRI.

| <u>Study</u> | <u>Gender F, M</u> | <u>Signs, symptoms, physical exam</u> | <u>MRI Findings</u> | <u>Proportion of patients with imaging-identified pulsatile tinnitus</u> | <u>Interventions/Outcomes</u> |
| --- | --- | --- | --- | --- | --- |
| Cortese 2021<sup>21</sup> | 8, 1 | Disappearance upon compression of ipsilateral internal jugular vein | -8 marginal sinus stenosis | 8/8 |  |
| Deuschl 2015<sup>23</sup> | 37,17 | Unspecified | -24 dAVF<br>-4 Paraganglioma<br>-2 AVM<br>-3 Extracranial Fistula<br>-2 Carotid-Cavernous Fistula<br>-1 ICA Stenosis<br>-1 ICA Aneurysm | 37/54 | N/A |
| Dietz 1994<sup>24</sup> | 31, 17 | -21 right PT<br>-4 bilateral PT<br>-18 left PT | Dural AVF (MRA best demonstrated transosseous channels and abnormal signals in the calvarium), paraganglioma (3/5 patients only had lesions identified on MR but not MRA. The other 2 patients were identified on both MR and MRA, with better identification on MRA), extracranial AVF, high jugular bulb (one out of the two patients was diagnosed only with MRA, not with MR, and the other was best demonstrated on MRA), jugular bulb diverticulum (1/1 best demonstrated on MRA), aberrant ICA, stenosis of ICA, carotid dissection, tortuous ICA, maxillary AVM | 28 | N/A |
| Lynch et al. 2021<sup>6</sup> | 186, 65 | Unspecified | -Arteriopathy<br>-Venopathy<br>-Middle/inner ear pathology<br>-Neoplasm | 49/80 |  |
| Sanchez 1998<sup>66</sup> | 15, 1 | -6 right PT<br>-5 left PT<br>-4 bilateral PT<br>-2 head PT | None identified by MRI. All identified by MRA, as specified in Table 5. | 13 | N/A |
| Shweel 2013<sup>41</sup> | 11, 16 | -right 13<br>-left 18 | DAVF (4), high jugular bulb (2), aneurysm of ICA (1), aberrant ICA (1), | 11 | N/A |
|  |  | -4 bilateral | vertebral artery hypoplasia (2),<br>glomus tumor (1) |  |  |
| Tsai et al. 2016<sup>59</sup> | Unspecified | -155 with PT | -25/28: suspicion for DAVF,<br>underwent further imaging with<br>MR/MRA or CT/CTA<br>-3/28 carotid stenosis, no further<br>imaging | 28/155 | N/A |
| Tao 2021<sup>49</sup> | Unspecified | - 69 left-sided pulsatile tinnitus<br>- 85 right-sided tinnitus<br>- 33 bilateral tinnitus<br>- 29 cases of migraine<br>- 54 cases of visual symptoms<br>- 102 cases of headaches<br>- 76 cases with symptoms<br>influenced by exercise or<br>activity<br>107 cases with symptoms influenced by<br>neck compression | -1 jugular bulb diverticulum<br>-1 large condylar veins<br>-3 prominent emissary vein<br>-1 venous sinus aneurysm<br>-3 venous sinus stenosis<br>-3 venous sinus stenosis and post<br>stenotic venous<br>aneurysm/diverticulum<br>-1 DAVF<br>-1 AVM | 14/66 | N/A |
| Sismanis 1990<sup>63</sup> | 30 total,<br>unspecified | Presented with tinnitus, suspicion for<br>benign intracranial hypertension. | All findings made on both CT and<br>MRI. 12 patients with empty sella, 4<br>patients with small ventricles, and<br>Chiari Type 1 malformation in one<br>patient. | 17/30 | Final diagnosis made based on<br>opening LP pressures |
| Deuschl 2015<sup>23</sup> | 37,17 | Unspecified | -24 dAVF<br>-4 Paraganglioma<br>-2 AVM<br>-3 Extracranial Fistula<br>-2 Carotid-Cavernous Fistula<br>-1 ICA Stenosis<br>-1 ICA Aneurysm | 37/54. Although DSA was<br>considered to be gold standard,<br>MRI was shown to be equivalent<br>in this study. | N/A |

**Supplemental Table 7.** DSA (including IVDSA)

| <u>Study</u> | <u>Gender F, M</u> | <u>Signs, symptoms, physical exam</u> | <u>DSA Findings</u> | <u>Proportion of patients with imaging-identified pulsatile tinnitus</u> | <u>Interventions/Outcomes</u> |
| --- | --- | --- | --- | --- | --- |
| Baomin 2014<sup>55</sup> | 44,2 | - 44 unilateral (34 right-sided, 12 left-sided)<br>- 2 bilateral<br>- Pulsatile tinnitus symptom reduced with head rotation or ipsilateral neck compression | -46/46 with transverse sigmoid sinus stenosis | 46/46 | -All patients failed an unspecified medical therapy.<br>-All patients underwent angioplasty and stenting and had complete resolution with no recurrent symptoms after 36 moth follow-up period |
| Carmody 1984<sup>58</sup> | 13,2 | - 15/15 pulsatile tinnitus<br>- 3/15 hearing loss<br>- 4/15 had a red mass in middle ear cavity<br>- 1/15 external otitis<br>- 2/15 bruit | -5/15 vascular mass<br>-1/15 frontal lobe AVM<br>-2/15 probable jugular vein thrombosis<br>-1/15 large jugular bulb | 9/15 | -4/15 patients with vascular mass underwent radiation and/or surgical resection<br>-1 case of AVM was treated with surgical resection |
| Lekovic 2021<sup>31</sup> | 13,2 | - 7/15 left-sided pulsatile tinnitus<br>- 5/15 right-sided pulsatile tinnitus<br>- 3/15 bilateral pulsatile<br>- 2/16 sensorineural hearing loss<br>- 5/16 had objective pulsatile tinnitus<br>- 4/16 had symptom reduction when compressing ipsilateral neck | -6 dAVF<br>-1 fibromuscular dysplasia<br>-1 Sigmoid Sinus diverticulum<br>-2 venous outflow obstruction with stenosis of ipsilateral TS sinus<br>-1 venous outflow obstruction at the skull base due to compression of the internal jugular vein by the enlarged styloid process. | 10/15 | -4 patients received embolization, with one patient also undergoing stenting<br>-1 patient had a craniotomy which was followed by onyx embolization for residual dAVF<br>-1 patient underwent decompression of the jugular vein at the skull base<br>-Of the 6 patients who underwent treatment, 5 had complete resolution |
| Sila 1987<sup>42</sup> | 15,5 | - 5 right-sided pulsatile tinnitus<br>- 10 left-sided pulsatile tinnitus<br>- 5 bilateral pulsatile tinnitus<br>- 14 patients had objective pulsatile tinnitus<br>- 14 patients had audible bruits<br>- 1 case of contralateral complete horner's syndrome<br>- 2 cases of papilledema<br>- 1 case of ipsilateral facial weakness | -1 left-sided carotid siphon stenosis<br>-1 left ICA minimal stenosis<br>-1 right ICE, left carotid siphon<br>-1 Bilateral carotid dissections<br>-1 pan arterial ectasia<br>-5 dural AVM of transverse sinus<br>-1 parietal AVM<br>-1 ICA occlusion | 14/20 | N/A |
| Deuschl 2015<sup>23</sup> | 37,17 | - Unspecified | -24 dAVF<br>-4 Paraganglioma<br>-2 AVM<br>-3 Extracranial Fistula<br>-2 Carotid-Cavernous Fistula<br>-1 ICA Stenosis<br>-1 ICA Aneurysm | 37/54. Although DSA was considered to be gold standard, MRI was shown to be equivalent in this study. | N/A |
| Shownkeen 2001<sup>40</sup> | 3, N/A | - 1 with dilated vein in posterior auricular region, found to have a dAVM<br>- 3 unilateral pulsatile tinnitus | -1 dAVF<br>-2 dAVM | 1/3 not diagnosed by MRI, needed further imaging with DSA<br>2/3 went directly to DSA for diagnosis |  |
| Ma 2015<sup>33</sup> | 2,1 | - 2 right-sided pulsatile tinnitus<br>- 1 left-sided pulsatile tinnitus<br>- 1 case of symptom reduction when compressing ipsilateral jugular vein | -2 dehiscence of sigmoid sinus wall and a mastoid emissary vein on the right side<br>-1 small diverticula of right sigmoid sinus and vascular malformation of right frontal-parietal lobe | 3/3 | -All 3 received treatment, and 2 had resolution of PT. -1 patient had a 70% reduction in pt symptoms following partial sigmoid sinus dehiscent wall reconstruction.<br>-The other patient experienced complete resolution of her symptoms after interventional therapy.<br>-The last patient had no change in her symptoms even after undergoing surgery for a mastoid emissary vein. |
| Tao 2021<sup>49</sup> | Unspecified | - 69 left-sided pulsatile tinnitus<br>- 85 right-sided tinnitus<br>- 33 bilateral tinnitus<br>- 29 cases of migraine<br>- 54 cases of visual symptoms<br>- 102 cases of headaches<br>- 76 cases with symptoms influenced by exercise or activity<br>- 107 cases with symptoms influenced by neck compression | -1 jugular bulb diverticulum<br>-1 large condylar veins<br>-3 prominent emissary vein<br>-1 venous sinus aneurysm<br>-3 venous sinus stenosis<br>-3 venous sinus stenosis and post stenotic venous aneurysm/ diverticulum<br>-1 DAVF<br>-1 AVM | 14/66 | N/A |

**Supplemental Table 8.**
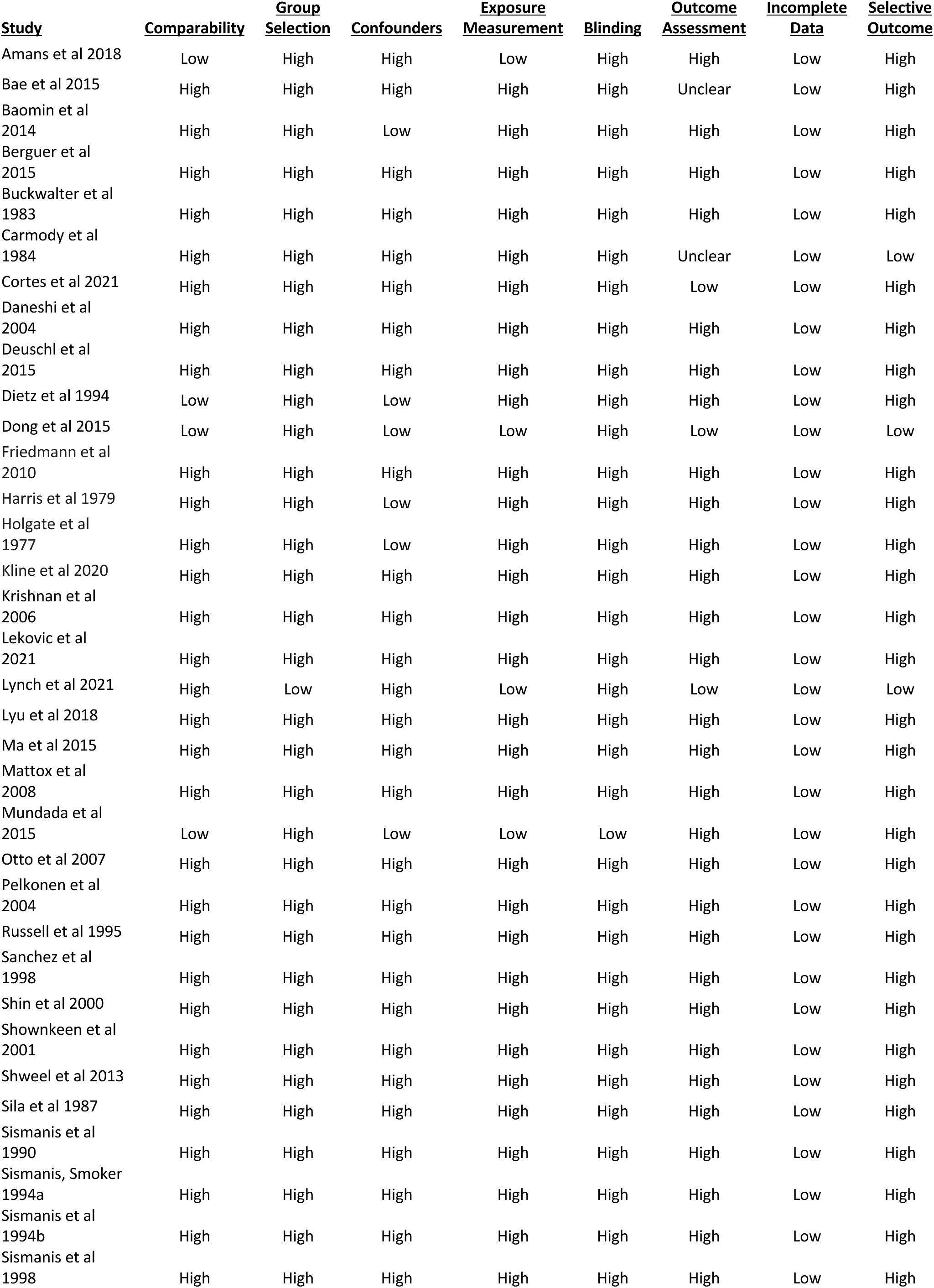
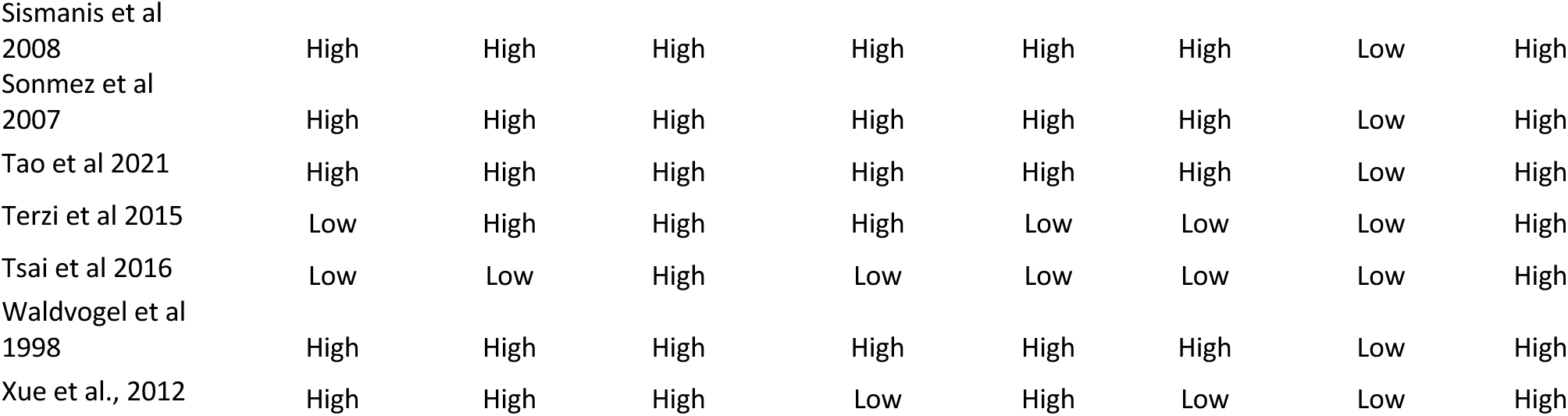
RoBANS 2 Scoring.

